# Antimicrobial resistance of *Staphylococcus* spp. from human specimens submitted to diagnostic laboratories in South Africa, 2012–2017

**DOI:** 10.1101/2024.07.07.24310040

**Authors:** Themba Titus Sigudu, Daniel Nenene Qekwana, James Wabwire Oguttu

## Abstract

**Background:** Antimicrobial drug resistance is of public health importance due to its potential to reduce treatment options and increase healthcare expenditure. There is, however, a paucity of studies that have examined antimicrobial resistance in countries with poor to moderate income. The present study examined the patterns and predictors of antimicrobial resistance in Staphylococcus isolates collected from humans at diagnostic laboratories in South Africa between 2012 and 2017.

**Method and materials:** A cross-sectional study design using retrospective data of 404 217 diagnostic laboratory records of staphylococcal isolates collected between 2012 and 2017 was adopted in this study. Isolates were assessed for antimicrobial drug resistance against 24 antimicrobials. Descriptive statistics, and binary logistic regression models were used to analyse the data. Significance was assessed at < 0.05.

**Results:** The highest resistance was observed against Cloxacillin (70.3%), while the lowest resistance was against Colistin (0.1%). A significant (p < 0.05) decreasing trend in AMR was observed over the study period, while a significant increasing temporal trend (p < 0.05) was observed for MDR over the same period. A Significant (p < 0.05) association was observed between specimen type, species of organism, and year of isolation with AMR outcome. Significant (p < 0.05) associations were observed between specimen type and season, with MDR.

**Discussion and recommendations:** The observed high levels of AMR and the increasing temporal trend in MDR is of public health concern. Clinicians should consider these findings when deciding on therapeutic options. Continued monitoring of AMR among *Staphylococcus* spp. and judicious use of antimicrobials in human medicine should be promoted.

## Introduction

An antimicrobial is an agent that kills microorganisms or stops their growth. But antimicrobial agents that act against bacteria are called “antibiotics”(Colgan *et al*., 2008). As antibiotics are used and in some cases, misused, resistance results (Sakwinska *et al*., 2011). Antimicrobial resistance (AMR) occurs when microorganisms become resistant to antimicrobial agents. This reduces the types of antibiotics that are available for treatment of infections. Causes of antibiotic resistance include the over-prescription of antibiotics, incorrect duration of treatment, lack of hygiene and poor sanitation, poor infection control practices in hospitals and clinics and the over-use of antibiotics in livestock and fish farming (Osimani and Clementi, 2016).

The development of antimicrobial agents has been one of the most important advancements in human and veterinary medicine over the last century (Oliveira and De Lencastre, 2002). However, the emergence of AMR agents in *Staphylococcus* spp. has become a significant public health threat as there are limited, or even sometimes no effective antimicrobial agents available to treat infections caused by these bacteria in both humans and domestic animals (Becker et al., 2015; Al Zahrani, 2011). Globally, AMR is estimated to be associated with over 700 000 deaths every year, a number which could rise to as high as 10 million in 2050 (Robinson and Enright, 2003).

The abundance of antimicrobials used in human and veterinary treatment has aided in the development of AMR (Huttner *et al*., 2013). The increased levels of AMR in pathogenic *Staphylococcus* spp., whether to a single agent or several antimicrobial classes, limits the capacity to treat illnesses successfully leading to increased morbidity (Reygaert, 2018). Moreover, there is evidence to suggest that AMR-related morbidity and mortality have increased in vulnerable communities (Hariyanto *et al*., 2022). The identification of resistance profiles of microorganism is therefore an important step in understanding AMR and is valuable in providing information to guide treatment options and to address the problem (Reygaert, 2018).

As reported by the World Health Organisation, the prevalence of resistance to first-line antimicrobials traditionally used to treat *Staphylococcus* infections has increased globally (WHO., 2022). However, this resistance is not confined to human medicine but is becoming more common in domestic species, particularly in equine medicine.

Given the potential of cross-transmission of some bacteria between humans and domestic species, identifying and describing the prevalence of AMR among domestic species has become even more crucial. In previous years, both the Centres for Disease Control and Prevention (CDC) and the United States Department of Agriculture (USDA) reported similar findings (Wolfe, Dunavan and Diamond, 2007). The findings of these studies revealed evidence of a possible zoonotic transfer of *Staphylococcus* bacteria and/or their genetic material between healthy humans and horses. Bramble *et al*. (2011) support the idea that resistant *Staphylococcus* infections in domestic animals may contribute to human-to-human transmission.

*Staphylococcus* species which exhibit multiple drug resistance (MDR) mechanisms, have been linked to life-threatening nosocomial infections, particularly in critically sick patients, posing significant therapeutic difficulties for physicians. Therefore, evaluating the prevalence of antibiotic-resistant *Staphylococcus* infections in humans is crucial not only for understanding the risk to the vulnerable but also for effectively providing information to guide efforts to develop infection prevention programs. Despite the fact that many studies have focused on methicillin-resistant *S. aureus* in humans, many other *Staphylococcus* species exhibiting antimicrobial resistance are also clinically significant for comprehending the epidemiology of AMR in humans and animals (Vanderhaeghen and Dewulf, 2017; Graveland *et al*., 2010; Weese and van Duijkeren, 2010; Tsubakishita *et al*., 2010).

Despite the widespread usage of antimicrobials in human and veterinary medicine in South Africa, the epidemiology of antimicrobial drug resistance of *Staphylococcus* spp. in South Africa has not received the attention it deserves. Therefore, the aim of this study was to investigate antimicrobial drug resistance among staphylococcal isolates from human samples submitted to diagnostic laboratories between 2012 and 2017, so as to improve on our understanding of burden of antimicrobial drug resistance among *Staphylococcus* spp. including the temporal trends of AMR and the predictors of AMR and MDR.

### 4.4 Materials and methods

#### 4.4.1 Study design and data extraction

A cross-sectional retrospective study design was implemented to realise the objectives of the present study. Records of 404 217 *Staphylococcus* isolates from 123 diagnostic laboratories countrywide collected over the period 2012 and 2017 were extracted from the National Health Laboratory Service (NHLS) electronic database. These laboratories service the public health sector hospitals and receive samples from all levels of healthcare service (district, regional, tertiary and central) in South Africa. Specimens submitted for microbiological analysis included skin, blood, urine, catheters (central venous catheter and haemodialysis), nasopharyngeal fluid and specimens from other body sites. For each isolate, data extracted from the NHLS database included a combination of demographic (age and sex), spatial (province of origin), clinical (species of organism and specimen type), temporal (season) as well as information on antimicrobial susceptibility profile (resistance or susceptible).

#### 4.4.2 Data management

The data were inspected for inconsistencies such as missing information, incorrect addresses and duplicate entries. No duplicates were identified, and no mixed infections were reported. The variable ‘age’ was re-categorised into the following 14 categories using the cohort-component method for population estimation produced by Statistics South Africa (StatsSA, 2020), 0–4 years, 5–9 years, 10–14 years, 15–19 years, 20–24 years, 25–29 years, 30–34 years, 35–39 years, 40–44 years, 45–49 years, 50–54 years, 55–59 years, 60–64 years and > 65 years (p. 25). Months were categorised into four seasons: autumn (March, April and May); winter (June, July and August); spring (September, October and November) and summer (December, January and February). The type of specimen was classified into the following five categories: skin, urinary, blood, nasopharyngeal fluid and ‘All other sites’. The resistance status variable was reclassified into a binary outcome, resistant or susceptible, by reclassifying isolates that were “intermediate” as resistant. Antimicrobial resistance (AMR) was defined as resistance to at least one antimicrobial class. While multi-drug resistance (MDR) was defined as resistance to three or more antimicrobial classes (Magiorakos *et al*., 2012).

#### 4.4.3 Data analysis

All data processing and analyses were performed using Stata Statistical Software (‘StataCorp, 2017). The number for resistant isolates and their corresponding 95% confidence intervals (95% CI) were computed and presented based on time, person and place. Annual changes in the number of *Staphylococcus* spp. were displayed using temporal graphs. Simple and multivariate binary logistic regression models were used to determine whether age, sex, year, province of origin, specimen type, *Staphylococcus* species and season were associated with AMR. The model was constructed in two stages. In the first phase, univariate binary logistic regression models with “AMR, (1 = Resistant, 0 = Susceptible)” as the outcome and each of the factors in Table 4 as explanatory variables were fitted to the data. The multivariate binary logistic regression model was fitted to the data using a manual backwards selection technique in the second stage. Confounding was assessed by comparing the change in parameter estimates of the variables in the model with and without the suspected confounding variable. A 20% change in the estimate of any of the parameter estimates in the model was interpreted as of the variable in question was a confounder, which was subsequently included in the final effects model. For each variable included in the final model, odds ratios and their 95% confidence intervals were calculated.

#### 4.4.4 Ethical considerations

Access to the NHLS database and patient information is restricted to laboratories staff working within the NHLS and can only be accessed at the premises of the NHLS. Thus, data extraction was carried out by the NHLS staff and de-identified data were provided to the researcher. Confidentiality and anonymity were always maintained by ensuring that patient personal information was not included in articles and reports. In addition, permission to use the data was obtained from the NHLS. Ethical approval was obtained from the University of South Africa, College of Agriculture & Environmental Sciences, Health Research and Animal Research Ethics Committees (Ref: 2018/ CAES/107). The data were kept safe from unauthorised access, accidental loss or destruction. Data in the form of softcopies were kept as encrypted files in computers and flash drives.

## Results

### Descriptive statistics

#### Isolates examined in the study

Table 1 shows that 44.4% of AMR isolates were isolated from male participants, while females contributed the rest (41.7%). The number of AMR isolates from individuals whose sex was unknown was comparatively low (13.9%). Similarly, the majority of MDR isolates were from males (56.7%), with females contributing only 39.5%. The number of MDR isolates from individuals with unspecified sex was also relatively low (3.8%) (Table 1).

**Table 1:** Distribution and Antimicrobial resistance of human *Staphylococcus* isolates by age and sex in South Africa, 2012-2017.

| Variable | Total isolates |  |  | AMR<sup>a</sup> isolates |  |  | MDR<sup>b</sup> isolates |  |  |
| --- | --- | --- | --- | --- | --- | --- | --- | --- | --- |
|  | *n* | % | 95%CI<sup>c</sup> | *n* | % | 95%CI<sup>c</sup> | *n* | % | 95%CI<sup>c</sup> |
| **Age groups** | **404217** |  |  | **219086** | **54.2** |  | **153602** |  |  |
| ≥65 | 22517 | 5.6 | 0.544-0.576 | 11392 | 5.2 | 0.504- 0.536 | 7321 | 4.8 | 0.046-0.049 |
| 60-64 | 2438 | 0.6 | 0.054-0.066 | 7449 | 3.4 | 0.324- 0.364 | 693 | 0.5 | 0.004-0.005 |
| 55-59 | 7049 | 1.7 | 0.164-0.176 | 3067 | 1.4 | 0.124- 0.156 | 2070 | 1.3 | 0.013-0.014 |
| 50-54 | 11770 | 2.9 | 0.284-0.304 | 8983 | 4.1 | 0.384- 0.436 | 3159 | 2.1 | 0.020-0.022 |
| 45-49 | 14962 | 3.7 | 0.366- 0.374 | 5258 | 2.4 | 0.224- 0.264 | 3516 | 2.3 | 0.022-0.024 |
| 40-44 | 19620 | 4.9 | 0.486- 0.494 | 11612 | 5.3 | 0.504- 0.564 | 4458 | 2.9 | 0.028-0.030 |
| 35-39 | 26031 | 6.4 | 0.636- 0.644 | 14679 | 6.7 | 0.646- 0.694 | 5047 | 3.3 | 0.032-0.034 |
| 30-34 | 33685 | 8.3 | 0.826- 0.834 | 17308 | 7.9 | 0.766- 0.812 | 9699 | 6.3 | 0.062-0.065 |
| 25-29 | 34788 | 8.6 | 0.856- 0.864 | 14241 | 6.5 | 0.626- 0.674 | 9198 | 6.0 | 0.058-0.062 |
| 20-24 | 25897 | 6.4 | 0.636- 0.644 | 12926 | 5.9 | 0.564- 0.616 | 6442 | 4.2 | 0.041-0.043 |
| 15-19 | 9610 | 2.4 | 0.236- 0.244 | 9202 | 4.2 | 0.364- 0.476 | 1957 | 1.3 | 0.012-0.014 |
| 10-14 | 6144 | 1.5 | 0.146- 0.154 | 6573 | 3.0 | 0.276- 0.324 | 3128 | 2.0 | 0.019-0.021 |
| 5-9 | 13953 | 3.5 | 0.346- 0.354 | 11173 | 5.1 | 0.476- 0.546 | 4756 | 3.1 | 0.030-0.032 |
| 0-4 | 117566 | 29.1 | 0.289- 0.291 | 81500 | 37.2 | 0.364- 0.386 | 74378 | 48.4 | 0.481-0.488 |
| Unknown | 58187 | 14.4 | 0.138- 0.150 | 29138 | 13.3 | 0.130- 0.136 | 17779 | 11.6 | 0.114-0.118 |
| **Sex** | **404 217** |  |  | **219086** |  |  | **153602** |  |  |
| Male | 210 858 | 52.2 | 0.126- 0.134 | 97274 | 44.4 | 0.439- 0.482 | 87092 | 56.7 | 0.565-0.570 |
| Female | 181 270 | 44.8 | 0.443- 0.453 | 91359 | 41.7 | 0.413- 0.420 | 60673 | 39.5 | 0.392-0.397 |
| Unknown | 12 089 | 3.0 | 0.276- 0.324 | 30453 | 13.9 | 0.137- 0.141 | 5837 | 3.8 | 0.037-0.039 |

Out of the 17 age groups represented in the data, with an additional “Unknown” category, the highest number of AMR isolates was observed in the 0-4 years age group (37.2%), followed by individuals with unspecified age (13.3%). The age group 55-59 years exhibited the least number of AMR isolates (1.4%) (Table 1).

Likewise, the age group 0-4 years old, exhibited the highest number of MDR isolates (48.4%). Individuals with unknown age contributed the second highest number of isolates (11.6%), with the elderly age group (60-64 years) contributing the least number of MDR isolates (0.5%) (Table 1).

Out of the total number of isolates, 65% and 50.2% of the isolates were AMR and MDR respectively. Within the CoPS group, *S. aureus* exhibited the highest number of isolates that were both AMR (59.9%) and MDR (90.1%) (Table 2). 2.6% and 2.5%, of the AMR isolates were *Staphylococcus intermedius* and *S. pseudintermedius* respectively (Table 2). Additionally, within the CoPS group, 6.0% and 3.9% of the MDR isolates were *S. intermedius* and *S. pseudintermedius* respectively (Table 2).

**Table 2:** Distribution and antimicrobial resistance profiles of human *Staphylococcus* isolates by species of organisms in South Africa, 2012-2017.

| Variable | Total isolates |  |  | AMR<sup>a</sup> isolates |  |  | MDR<sup>b</sup> isolates |  |  |
| --- | --- | --- | --- | --- | --- | --- | --- | --- | --- |
|  | *n* | % | 95%CI<sup>c</sup> | *n* | % | 95%CI<sup>c</sup> | *n* | % | 95%CI<sup>c</sup> |
| **Staph species** | **404217** |  |  | **219086** |  |  | **153602** |  |  |
| **CoPS<sup>d</sup>** | **284973** | **70.5** |  | **142406** | **65.0** |  | **77108** | **50.2** |  |
| *S. aureus* | 270421 | 66.9 | 0.665- 0.672 | 131233 | 59.9 | 0.598- 0.600 | 69474 | 90.1 | 0.896 - 0.906 |
| *S. intermedius* | 8084 | 2.0 | 0.188- 0.212 | 5696 | 2.6 | 0.025- 0.026 | 4168 | 6.0 | 0.562 - 0.638 |
| *S. pseudintermedius* | 6468 | 1.6 | 0.148- 0.172 | 5477 | 2.5 | 0.024- 0.025 | 163 | 3.9 | 0.362 - 0.418 |
| **CoNS<sup>e</sup>** | **71546** | **17.7** |  | **50390** | **23** |  | **47309** | **30.8** |  |
| *S. epidermidis* | 45272 | 11.2 | 0.109-0.144 | 23004 | 10.5 | 0.836- 0.846 | 34678 | 73.3 | 0.729 - 0.737 |
| *S. haemolyticus* | 14552 | 3.6 | 0.336- 0.376 | 9640 | 4.4 | 0.434- 0.448 | 5583 | 16.1 | 0.156 - 0.166 |
| *S. hominis* | 7276 | 1.8 | 0.168- 0.192 | 11393 | 5.3 | 0.522- 0.538 | 402 | 7.2 | 0.682 - 0.758 |
| *S. saprophyticus* | 4446 | 1.1 | 0.108- 0.112 | 6353 | 2.9 | 0.286- 0.294 | 14 | 3.5 | 0.328 - 0.372 |
| **CoPS/CoNS<sup>f</sup>** | **47698** | **11.8** |  | **26290** | **12.0** |  | **29185** | **19.0** |  |
| Unspeciated *Staphylococcus* | 36784 | 9.1 | 0.902- 0.920 | 15117 | 6.9 | 0.686- 0.694 | 25945 | 88.9 | 0.884 - 0.894 |
| *S. schleiferi* | 5659 | 1.4 | 0.132- 0.148 | 5258 | 2.4 | 0.234- 0.244 | 1219 | 4.7 | 0.432 - 0.508 |
| *S. hyicus* | 5255 | 1.3 | 0.126- 0.134 | 5915 | 2.7 | 0.264- 0.274 | 78 | 6.4 | 0.602 - 0.678 |

The isolates belonging to the CoNS group constituted 23 and 30.8% of the total AMR and MDR isolates respectively. But within the CoNS group, *S. epidermidis*, contributed the highest number isolates that were AMR (11.2%) and MDR (73.3%). This was followed by *S hominis* which made up 5.3% of the AMR, while *S. haemolyticus* exhibited the second highest MDR isolates (16.1%) (Table 2). *Staphylococcus saprophyticus* contributed the least number of AMR (2.9%) and MDR (3.5%) (Table 2).

Up to 12 and 19% of the total isolates that were AMR and MDR respectively, belonged to the Coagulase-variable group. Within the coagulase-variable group, the Unspecified *Staphylococcus* species exhibited the highest number of AMR (6.9%) and MDR (88.9%) isolates, and *S. hyicus* contributed the second highest number of isolates that were AMR (2.7%) and MDR (6.4%). *Staphylococcus schleiferi* contributed the least number of AMR (2.4%) and MDR (4.7%) isolates in the data (Table 2).

### Resistance observed against different antimicrobial agents

This study evaluated a total of 24 antimicrobial agents, which were categorised into 9 distinct classes as shown in Table 3. Of these, 33.9% isolates showed AMR against Beta-lactams and 36.9% were involved in MDR combinations. Within the Beta-lactams, the highest levels of AMR was against Cloxacillin (70.3%), and likewise Cloxacillin was involved in the highest number of MDR combinations (74.2%). The second-highest level of AMR and MDR combinations was observed against Penicillin (14.6% and 13.7% respectively). Among the Beta-lactams, the lowest AMR was against Ertapenem (0.0%), and likewise, Ertapenem was involved in the least number of the MDR (0.1%) combinations.

**Table 3:** Distribution of antimicrobial resistance categorised by antimicrobial class among human *Staphylococcus* samples submitted to diagnostic laboratories in South Africa, 2012 – 2017.

|  | AMR<sup>a</sup> isolates |  |  | MDR<sup>b</sup> isolates |  |  |
| --- | --- | --- | --- | --- | --- | --- |
|  | *n* | % | 95%CI<sup>c</sup> | *n* | % | 95%CI<sup>c</sup> |
| **Beta-lactams:** | **136862** | **33.9** |  | **106917** | **36.9** |  |
| Ceftazidime | 411 | 0.3 | 0.003-0.004 | 214 | 0.2 | 0.18 - 0.22 |
| Amoxicillin | 3832 | 2.8 | 0.027-0.029 | 1711 | 1.6 | 1.48 - 1.72 |
| Ampicillin | 3148 | 2.3 | 0.022-0.024 | 2352 | 2.2 | 2.08 - 2.32 |
| Cloxacillin | 96147 | 70.3 | 0.701-0.706 | 79332 | 74.2 | 73.96 - 74.44 |
| Penicillin | 19982 | 14.6 | 0.144-0.148 | 14755 | 13.7 | 13.68 - 13.92 |
| Piperacillin | 274 | 0.2 | 0.002-0.003 | 107 | 0.1 | 0.08 - 0.12 |
| Cefazolin | 274 | 0.2 | 0.001-0.003 | 107 | 0.1 | 0.08 - 0.12 |
| Cefepime | 411 | 0.3 | 0.002-0.003 | 214 | 0.2 | 0.18 - 0.22 |
| Cefotaxime | 821 | 0.6 | 0.005-0.006 | 428 | 0.4 | 0.38 - 0.42 |
| Cefoxitin | 10128 | 7.4 | 0.073-0.075 | 6736 | 6.3 | 6.18 - 6.42 |
| Cefuroxime | 958 | 0.7 | 0.007-0.008 | 642 | 0.6 | 0.58 - 0.64 |
| Ertapenem | 67 | 0.0 | 0.000-0.000 | 100 | 0.1 | 0.00 - 0.02 |
| Imipenem | 274 | 0.2 | 0.001-0.002 | 214 | 0.2 | 0.18 - 0.22 |
| Meropenem | 137 | 0.1 | 0.001-0.001 | 107 | 0.1 | 0.08 - 0.12 |
| **Aminoglycosides:** | **53081** | **13.1** |  | **40494** | **14.1** |  |
| Gentamicin | 51655 | 97.3 | 0.992-0.993 | 37457 | 92.5 | 0.991-0.994 |
| Amikacin | 657 | 1.2 | 0.001-0.003 | 1701 | 4.2 | 0.004-0.005 |
| Streptomycin | 330 | 0.6 | 0.000-0.001 | 486 | 1.2 | 0.000-0.001 |
| Tobramycin | 439 | 0.8 | 0.002-0.009 | 850 | 2.1 | 0.002-0.003 |
| **Sulfonamides:** |  |  |  |  |  |  |
| Co-trimoxazole | 15498 | 3.7 | 0.037-0.038 | 12601 | 4.3 | 0.035-0.036 |
| **Tetracyclines:** |  |  |  |  |  |  |
| Tetracycline | 46311 | 12.1 | 0.120-0.122 | 31721 | 10.9 | 0.108-0.110 |
| **Macrolides:** |  |  |  |  |  |  |
| Erythromycin | 85591 | 21.2 | 0.211-0.213 | 56160 | 19.4 | 0.246-0.249 |
| **Lincosamides:** |  |  |  |  |  |  |
| Clindamycin | 56036 | 15.5 | 0.155-0.156 | 34759 | 12.1 | 0.190-0.192 |
| **Phenicol:** |  |  |  |  |  |  |
| Chloramphenicol | 10490 | 2.5 | 0.024-0.025 | 7159 | 2.5 | 0.027-0.028 |
| **Polymyxins:** |  |  |  |  |  |  |
| Colistin | 348 | 0.1 | 0.001-0.001 | 193 | 0.1 | 0.000-0.001 |

A total of 13.1% isolates exhibited AMR against aminoglycosides and 14.1% were involved in MDR combinations. The overwhelming majority of isolates that exhibited AMR against aminoglycosides, were resistant against Gentamicin (97.3%). Likewise, Gentamicin (92.5%) was involved in the overwhelming majority of MDR combinations. The second highest level of resistance among isolates that were resistant against aminoglycosides, were resistant against amikacin (1.2%), and the same amikacin was involved in 4.2% of the MDR combinations. Meanwhile, among aminoglycosides, the lowest level of resistance was observed against Streptomycin (0.6%), and the same antimicrobial drug was also involved in the lowest MDR combinations (1.2%).

Within the sulfonamides class, Co-trimoxazole was the only antimicrobial agent assessed, and only 3.7% of the isolates exhibited AMR against Co-trimoxazole, and the same drug was involved in 4.3% MDR combinations. Tetracycline was the only antimicrobial agent evaluated within the tetracyclines class, only 12.1% of the isolates exhibited AMR against Tetracycline, and similarly Tetracycline was involved in 10.9% MDR combinations.

In the macrolides class, Erythromycin was the only antimicrobial agent assessed and 21.2% of the isolates exhibited AMR against Erythromycin. Furthermore, Erythromycin was implicated in 19.4% of MDR combinations.

Clindamycin, being the only agent evaluated within the Lincosamides class, only 15.5%% of the isolates exhibited AMR against Clindamycin, and it was involved in 12.1% MDR combinations. Among the Phenicols, Chloramphenicol was the only agent examined, and only 2.5% of the isolates showed AMR against Chloramphenicol, and the same drug was involved in 2.5% MDR combinations. Of the Polymyxins examined, Colistin was the only antimicrobial agent analysed, and only 0.1% of the isolates displayed AMR against Colistin, and in addition, the same drug was involved in only 0.1% MDR combinations.

### Temporal trends

The annual trends for AMR and MDR over the study period, 2012 to 2017, are presented in Figure 1. There were fluctuations in the numbers of isolates that were AMR and MDR over the six-year period. For example, in 2013, a minor decrease in AMR from 78% in 2012 to 71% in 2013 was observed. However, over the same period, there was an increasing trend in MDR from 23.0% in 2012 to 26% in 2013.

**Figure 1:**
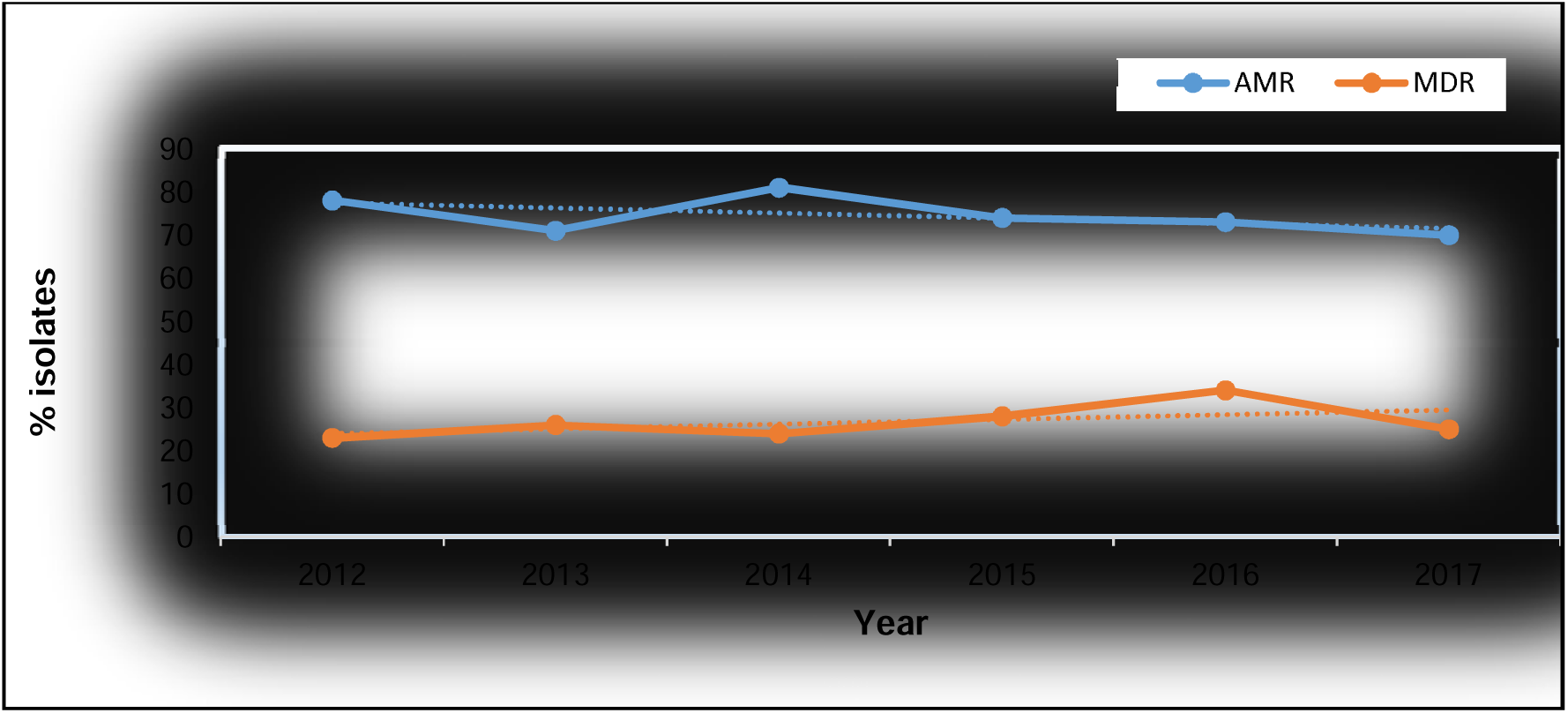
Annual temporal distribution of AMR and MDR from human Staphylococcus samples submitted to diagnostic laboratories in South Africa, 2012 - 2017

The following year, 2014, saw the highest percentage (81%) of AMR isolates over the entire six-year period. However, over the same period, the number of MDR percentage dropped to 24%. From 2014 to 2015, there was a visible decline in AMR, while the number of MDR slightly increased to 28%. From 2015 to 2016, the number of AMR isolates were relatively stable at 73%. But over the same period, the number of MDR isolates peaked at 34%, which was the highest over the six-year period. From 2016 to 2017, there was a decrease in the number of both AMR (70%) and MDR (25%) isolates recorded (Figure 1).

## Inferential statistics

### Predictors for antimicrobial drug resistance

*Staphylococcus aureus* had significantly higher odds (Adjusted Odds Ratio (AOR) = 2.6; 95% CI: 1.80-2.80) of being AMR as compared to *S. pseudintermedius* (reference category). On the other hand, *S intermedius* had lower odds (AOR=0.5; 95% CI: 0.30-0.90) of being AMR compared to the reference category (Table 4).

**TABLE 4:** Predictors of antimicrobial resistance among *Staphylococcus* from human samples submitted to diagnostic laboratories in South Africa, 2012–2017.

| Variable | OR | AMR<br>95%CI | P-Vale |
| --- | --- | --- | --- |
| Staph. Species |  |  |  |
| *S. aureus* | 2.6 | 1.80-2.80 | < 0.001 |
| *S. intermedius* | 0.5 | 0.30- 0.90 | < 0.001 |
| *S. pseudintermedius* | Reference | Reference | Reference |
| Specimen source |  |  |  |
| Skin | 1.6 | 1.20-2.10 | 0.243 |
| Urinary | 0.8 | 0.60-1.60 | 0.384 |
| Blood | 0.4 | 0.20-2.20 | 0.067 |
| Nasopharyngeal fluid | 0.9 | 0.60- 1.80 | 0.672 |
| All other sites | Reference | Reference | Reference |
| Year | 0.97 | 0.94-1.10 | 0.022 |

**TABLE 5:** Predictors of multidrug resistance among *Staphylococcus* from different human samples submitted to diagnostic laboratories in South Africa, 2012–2017.

| Variable | MDR |  |  |
| --- | --- | --- | --- |
|  | OR | 95%CI | P-Vale |
| Specimen source |  |  | < 0.001 |
| Skin | 2.7 | 0.80-11.60 | 0.246 |
| Urinary | 6.0 | 2.20-17.60 | 0.407 |
| Blood | 13.0 | 3.40-55.20 | 0.001 |
| Nasopharyngeal fluid | 2.4 | 0.80-7.80 | 0.265 |
| Other sites | - | - | - |
| Season |  |  | 0.022 |
| Autumn | 0.9 | 0.80-1.30 | 0.152 |
| Winter | 1.8 | 1.20-2.70 | 0.045 |
| Spring | 1.6 | 0.40-4.60 | 0.355 |
| Summer | - | - | - |

The isolates from skin specimens had higher odds (AOR=1.6, 95% CI: 1.20-2.10) of being AMR compared to the reference category (All other sites). However, this association was not statistically significant (p = 0.243). The isolates from the urinary specimens compared to the reference category, had lower odds (AOR= 0.8; 95% CI: 0.60-1.60) of being AMR. However, this association was also not statistically significant (p = 0.384). The isolates from the blood specimens compared to the reference category, had lower odds (AOR=0.4; 95% CI: 0.20-2.20) of being AMR. However, this association was marginally statistically significant (p = 0.067). The isolates from the nasopharyngeal fluid specimens compared to the reference category had lower odds (AOR=0.9; 95% CI: 0.60-1.80) of being AMR. However, the association also did not reach significance (p = 0.672).

### Predictors for multi drug resistance

The odds ratio of 2.7 (95% CI: 0.80-11.60) suggests an increased odds of occurrence of MDR among isolates from the skin specimens as compared to the reference category (Other sites). However, this association was not statistically significant (p = 0.246). The odds ratio of 6.0 (95% CI: 2.20-17.60) taphylococcal isolates from the urinary specimens had higher odd of occurrence of MDR compared to the isolates from the reference category. This association was not statistically significant (p = 0.407). Isolates from blood specimens had significantly (p = 0.001) higher odds (AOR= 13.0 (95% CI: 3.40-55.20) of being MDR compared to the reference category. isolates from the nasopharyngeal fluid specimens had higher odds (AOR=2.4; 95% CI: 0.80-7.80) of being MDR compared to the reference category. However, this association was not statistically significant (p = 0.265).

The overall p-value for the association between season and MDR was statistically significant (p = 0.022). Isolates recovered in autumn had higher odds (AOR= 0.9, 95% CI: 0.80-1.30) of being MDR compared to the isolates obtained in summer (reference category). This association was not statistically significant (p = 0.152). The isolates obtained during winter had significantly (p = 0.045) higher odds (AOR=1.8; 95% CI: 1.20-2.70) as compared to the reference category. Although not statistically significant (p = 0.355), the isolates obtained during spring had higher odds (AOR=6; 95% CI: 0.40-4.60) of being MDR as compared to the reference category.

## Discussion

The results of this study hold significant implications for understanding AMR among *Staphylococcus* isolates in South Africa over the study period (2012 to 2017). Firstly, the observed higher number of AMR isolates among males compared to females underscores the importance of considering potential sex-based differences in the risk factors associated with acquiring and transmitting resistant bacterial strains. This suggests that there may be gender-specific factors influencing the prevalence of AMR, which warrant further investigation. Furthermore, the alignment of these findings with a previous multicentre investigation in Asmara, Eritrea, suggests a potential regional consistency in the gender distribution of AMR *Staphylococcus* isolates (Khanna *et al*., 2008). However, the contrasting pattern observed in another study conducted in Myanmar highlights the complexity of AMR epidemiology and the need for context-specific analysis (Soe *et al*., 2021).

The findings from this study demonstrated the distribution of AMR and MDR isolates across different age groups, shedding light on potential of age in influencing susceptibility to infections and patterns of resistance. Available evidence shows that children aged 0-4 years are more vulnerability to bacterial infections, likely due to their developing immune systems and close interactions in day-care or nursery settings (Bloomfield *et al*., 2012; Sigudu, Oguttu and Qekwana, 2023). Additionally, the high prevalence of isolates in this age group may reflect increased healthcare-seeking behaviours for young children (Nielsen *et al*., 2012).

The concerning trend of high numbers of both AMR and MDR isolates among children aged 0-4 years underscores the urgent need for targeted interventions in this vulnerable population. Factors such as previous antibiotic exposure, environmental influences, and transmission dynamics within households or childcare settings may contribute to this high prevalence, necessitating multifaceted approaches including improved antibiotic stewardship and infection control measures (Waterlow *et al*., 2024). Efforts to enhance data collection and reporting for individuals with unspecified ages could provide further insights into the patterns of AMR and MDR isolates across populations.

Moreover, although the age group 60-64 years demonstrated lower numbers of AMR and MDR isolates, it is crucial to consider variations within the broader elderly population, as factors such as immune function, healthcare exposure, and antibiotic usage can influence resistance patterns (Baquero *et al*., 2021; Kardaś-Słoma *et al*., 2011). However, the low numbers of AMR and MDR among this age group could be explained the by the low carriage of *Staphylococcus* among the age group 60-64 years reported in previous studies (Hayward et al., 2021). These findings, suggest potential differences in health-seeking behaviours, age-specific immunity, or reduced exposure to sources of infection in this older demographic.

The results of the examination of 32 distinct *Staphylococcus* species provide crucial insights into the prevalence and distribution of AMR across different categories of coagulase, highlighting significant implications for public health and clinical practice (Abdullahi *et al*., 2023). The high numbers of AMR and MDR observed among the CoPS group, particularly *S. aureus* indicate the urgent need for effective strategies to combat resistance in this clinically significant pathogen, as its resistance can lead to high morbidity, mortality, and healthcare costs (Denissen *et al*., 2022). Furthermore, the presence of AMR in other CoPS species like *S. intermedius* and *S. pseudintermedius*, albeit in smaller numbers, reinforces the importance of surveillance and intervention measures targeting a broad spectrum of *Staphylococcus* species (Saputra, 2018).

Among CoNS, the high prevalence of AMR in species like *Staphylococcus epidermidis* highlights the challenge posed by resistance in non-pathogenic strains, which can still cause infections in vulnerable populations and serve as reservoirs for resistance genes (França *et al*., 2021). The high numbers of AMR observed across both pathogenic CoPS strains and non-pathogenic CoNS strains emphasise the urgent need for comprehensive strategies to address antimicrobial resistance in *Staphylococcus* species. Such strategies should encompass enhanced antibiotic stewardship, infection control measures, and research into alternative treatment options to mitigate the impact of resistance on public health and clinical outcomes (Owens, 2008).

The analysis of antimicrobial agents conducted in this study provides critical insights into the observed resistance patterns, with the highest levels of resistance observed against Beta-lactams. Notably, the high levels of resistance were observed against Cloxacillin, a narrow-spectrum antibiotic which primarily targets Gram-positive bacteria, demonstrated the highest rates of both antimicrobial resistance (AMR) and multidrug resistance (MDR) among Beta-lactams. This suggests prevalent resistance within the examined population (Bru and Garraffo, 2012). This finding underscores the necessity for close monitoring and prudent use of Cloxacillin to mitigate the spread of resistance (Leong *et al*., 2018). Penicillin, another Beta-lactam antibiotic, exhibited the second-highest rates of AMR and MDR. The broader spectrum of activity of Penicillin, targeting a wider range of bacteria, could contribute to the observed resistance levels (Lambert, 2005). In contrast, Ceftazidime, a third-generation cephalosporin, exhibited the lowest rates of AMR and MDR among the Beta-lactams. This suggests that Ceftazidime may remain an effective treatment option for a significant number of bacterial isolates, particularly where resistance to other Beta-lactams is present (Tamma and Rodriguez-Baňo, 2017; Lagacé-Wiens, Walkty and Karlowsky, 2014).

Among the aminoglycosides class, Gentamicin exhibited alarmingly high resistance rates, with nearly all isolates showing both AMR and MDR. This high level of resistance suggests that Gentamicin may have limited effectiveness against many bacterial infections due to widespread resistance (Gad, Mohamed and Ashour, 2011). Tobramycin demonstrated the lowest AMR and MDR rates among aminoglycosides, suggesting it may be a more suitable treatment option in some cases (Horcajada *et al*., 2019).

Co-trimoxazole, the only sulfonamides assessed, showed modest rates for both AMR and MDR. This finding is intriguing because sulfonamides, including co-trimoxazole, have historically been associated with high levels of resistance due to their extensive use and mechanisms that bacteria can develop to counteract their effects (European Centre for Disease Prevention and Control, 2020, 2020). While modest rates for both AMR and MDR against co-trimoxazole were unexpected, given the historical trends with sulfonamides, several factors could explain this observation, including antibiotic stewardship efforts, combination therapy effects, limited usage, population dynamics, geographical variability, and the ongoing evolution of resistance mechanisms (Scaglione *et al*., 2022). Various studies have reported on the level of resistance against sulfonamides like co-trimoxazole. Many of these studies have highlighted a concerning trend of increasing resistance over time, particularly in regions where these antibiotics are commonly prescribed and where there may be inadequate regulations on their use (Ayukekbong, Ntemgwa and Atabe, 2017).

Tetracycline, the sole representative of the tetracyclines class, displayed notable resistance levels. In view of this, resistance to this antibiotic should be monitored closely (Alhamami *et al*., 2022). The notable resistance levels of tetracycline could be attributed to various factors, including mechanisms of resistance and implications for human use. Research indicates that bacteria can develop resistance to tetracycline through mechanisms such as efflux pumps, ribosomal protection proteins, and enzymatic inactivation (Chopra and Roberts, 2001). These mechanisms enable bacteria to survive and proliferate despite tetracycline exposure. Epidemiological studies highlight the prevalence of tetracycline resistance among various bacterial pathogens, underscoring the importance of monitoring resistance patterns (Livermore, 2000). This surveillance is crucial for guiding antibiotic treatment strategies and public health policies aimed at combating resistance. Erythromycin, the only macrolide examined, exhibited significant resistance rates for both AMR and MDR, indicating a notable level of resistance that could impact its effectiveness in treating infections (Food and Authority, 2015). The high resistance rate observed against erythromycin is a multifaceted issue influenced by several key factors identified in several studies. Firstly, selective pressure from the excessive and inappropriate use of antibiotics, including erythromycin, favours bacteria with pre-existing resistance or those acquiring resistance mutations through natural selection. This selective pressure promotes the survival and proliferation of resistant bacterial strains, thereby diminishing erythromycin’s efficacy in treating infections (Cantón and Morosini, 2011). Secondly, genetic mechanisms significantly contribute to erythromycin resistance, with studies demonstrating how bacterial acquisition of methylase-encoding genes can alter ribosomal target sites, reducing the antibiotic’s ability to inhibit bacterial protein synthesis effectively (Desjardins *et al*., 2004). Moreover, concerns about cross-resistance are prominent, as resistance mechanisms developed against erythromycin may confer resistance to other antibiotics within the macrolide class and occasionally across different antibiotic classes. This complicates treatment strategies and underscores the importance of prudent antibiotic use, alongside the development of innovative antimicrobial agents with novel mechanisms of action (Muteeb *et al*., 2023).

The observed high levels of AMR and involvement in MDR combinations against clindamycin, a member of the lincosamides class, can be attributed to several factors reported in various studies that have identified various lincosamide-modifying enzymes (LMEs) that inactivate clindamycin. For example, studies by Mitcheltree, (2018) and Fiebelkorn *et al*. (2003) demonstrated the presence of LMEs in clinical isolates of *Staphylococcus aureus* and *Enterococcus* spp., contributing to resistance against clindamycin. Mutations in the ribosomal binding site have been documented in studies, such as those by Roberts, Pea and Lipman (2013), showing how alterations can reduce clindamycin binding affinity, thereby decreasing its effectiveness. Studies, including those by Malbruny *et al*. (2011), have demonstrated cross-resistance between clindamycin and other lincosamide antibiotics due to shared resistance mechanisms. This cross-resistance complicates treatment options when faced with resistant strains.

Chloramphenicol, a member of the Phenicol class of antimicrobials, displayed low AMR and MDR rates, suggesting it may still be a viable option for certain infections. The lower resistance rates observed for chloramphenicol can be attributed to several factors based on studies and epidemiological data. Chloramphenicol acts by inhibiting bacterial protein synthesis through binding to the 50S ribosomal subunit, similar to clindamycin. However, the specific mechanisms of resistance differ. Chloramphenicol resistance often involves enzymatic inactivation by chloramphenicol acetyltransferases (CATs) or efflux pumps, Studies like the one by Schwarz *et al*. (2004) have highlighted these distinct resistance mechanisms. Chloramphenicol, once widely used, has seen reduced clinical use due to toxicity concerns (e.g., aplastic anemia) but remains effective in some settings where resistance rates are lower (Dinos *et al*., 2016). Resistance rates against chloramphenicol can also vary geographically and over time. Studies have shown that chloramphenicol may retain efficacy in certain regions or against specific pathogens where resistance to other antibiotics, is more prevalent (Shafran, 1990). For example, surveillance studies conducted by national health agencies or international organisations often report varying resistance patterns that influence treatment guidelines.

Colistin, the only Polymyxins assessed, showed minimal AMR and no MDR, indicating its potential as a viable treatment option for certain bacterial infections (Pacheco *et al*., 2019). Colistin acts by disrupting the bacterial cell membrane, leading to leakage of intracellular contents and ultimately bacterial cell death. This mechanism differs significantly from many other classes of antibiotics, which may contribute to its unique profile of resistance (L. Kakoullis, E. Papachristodoulou, 2021). Resistance to Colistin is primarily mediated by chromosomal mutations in genes such as those encoding the lipid A biosynthesis pathway (e.g., pmrAB and phoPQ systems). Acquisition of plasmid-mediated resistance genes (e.g., mcr genes) has been reported but remains relatively uncommon compared to other antibiotic classes (Meng, Li and Yao, 2022). Colistin has historically been reserved as a last-line treatment option, particularly for infections caused by multidrug-resistant Gram-negative bacteria such as Pseudomonas aeruginosa, Acinetobacter baumannii, and carbapenem-resistant Enterobacteriaceae (CRE). Its limited use and stringent clinical indications may contribute to the observed low levels of resistance (Beceiro, Tomás and Bou, 2013).

The annual trends in AMR and MDR observed in this study from 2012 to 2017 indicate fluctuating patterns, emphasizing the dynamic nature of resistance development and the need for continuous monitoring. In 2013, a slight decrease in AMR rate was observed compared to the previous year (2012), accompanied by a notable increase in the rate of MDR. This shift may reflect changes in antibiotic usage patterns or the emergence of new resistant strains during that period (Barbosa and Levy, 2000). In 2014, the highest AMR rate was recorded during the six-year period, while MDR rate dropped. This highlights the importance of ongoing surveillance, as resistance rates can change significantly from year to year (Aidara-Kane, 2012). In 2015, a decline in AMR rate was accompanied by a slight increase in the rate of MDR. This pattern suggests that while overall resistance to individual antibiotics may have decreased, the number of isolates resistant to multiple drugs can remain a concern (Yaovi *et al*., 2022). In 2016, the AMR rate remained relatively stable, while MDR reached its peak throughout the six-year period. This indicates that despite the overall resistance levels staying constant, the number of isolates MDR increased, and this poses a significant challenge for treatment options (Winkler *et al*., 2015).

The results of the present study provides valuable insights into the predictors of AMR among *Staphylococcus* species, based on specimen source and year. *Staphylococcus aureus* showed significantly higher odds of being AMR compared to *S. pseudintermedius*, suggesting that certain species within the *Staphylococcus* genus may inherently possess greater resistance mechanisms or are more prone to acquiring resistance (Bitrus *et al*., 2018). Conversely, *S. intermedius* exhibited lower odds of AMR compared to the reference category, indicating potential differences in susceptibility profiles among *Staphylococcus* species (Diekema *et al*., 2001).

Specimen source improved to be a significant predictor of AMR. For example, isolates from skin specimens had higher odds of being AMR compared to isolates from other sites, highlighting the importance of considering the site of infection when assessing resistance patterns (Schmidt *et al*., 2014). Similarly, isolates from the urinary specimens had lower odds of AMR, albeit not statistically significant, suggesting potential differences in resistance prevalence across different types of infections (Waterlow *et al*., 2024). The marginally statistically significant association observed between isolates from blood specimens compared to isolates from specimen designated “other types” suggests that Staphylococcus species from different sites may exhibit different resistance patterns (Ombelet *et al*., 2022).

The finding that seasonality was a significant predictor of MDR among staphylococcal isolates, with higher occurrence in winter compared to summer, is a reflection of several interconnected factors that have been observed in studies examining antibiotic resistance patterns. For example the cold weather during winter months can create conditions favouring the survival and transmission of certain bacterial pathogens (Hellberg and Chu, 2016). This includes staphylococci, which may thrive in colder temperatures or have increased survival rates on surfaces during winter. In addition, during winter, people tend to spend more time indoors in close proximity, facilitating the spread of bacteria and potentially contributing to higher rates of MDR infections in healthcare and community settings (Ben Maamar, Hu and Hartmann, 2020). Furthermore, winter months often coincide with peaks in respiratory infections and other illnesses, leading to higher healthcare utilization. This may contribute to increased exposure to antibiotics and selection pressure for multidrug-resistant strains among staphylococcal infections (Steinberg et al., 2013). Moreover, seasonal variations in immune function and vitamin D levels among individuals may affect susceptibility to infections and response to antibiotic treatment (Ismailova and White, 2022). Lower immune responses during winter could potentially exacerbate the severity and persistence of staphylococcal infections, increasing the likelihood of multidrug resistance (Gehrke, Giai and Gómez, 2023).

## Limitations of the Study

The utilisation of diagnostic laboratory data stands as a notable strength of the study, offering a substantial dataset for the analysis of AMR patterns. This approach provides a robust foundation for understanding the prevalence and distribution of resistant bacterial strains. Moreover, the focus on South Africa allows for the generation of region-specific findings, offering valuable insights into the epidemiology of AMR within the context of the country’s healthcare landscape. However, the study is subject to several limitations that warrant consideration. Firstly, the reliance on diagnostic laboratory data may introduce selection bias, as the sampled population may not be representative of the broader population. This limitation could potentially skew the study’s findings and limit their generalizability to other settings. Furthermore, the geographical scope limited to South Africa may restrict the applicability of the study’s findings to regions with different healthcare systems, antibiotic usage patterns, and population demographics.

## Conclusions

The results of this study provide significant insights into AMR in *Staphylococcus* isolates in South Africa. Firstly, the observed higher number of AMR isolates among males compared to females suggests potential sex-based differences in the risk factors associated with acquiring and transmitting resistant bacterial strains. This underscores the importance of further investigating gender-specific factors influencing AMR prevalence. Furthermore, disparities observed in the present and other studies highlight the complexity of AMR epidemiology and the need for context-specific analysis.

The low number of AMR and MDR isolates in the 60-64 age group on one hand, and the high numbers of both AMR and MDR isolates among young children, emphasizes the urgent need for targeted interventions to reduce the prevalence of AMR and MDR. The analysis of distinct *Staphylococcus* species provides crucial insights into the prevalence and distribution of AMR across different coagulase categories, highlighting significant implications for public health and clinical practice. The dominance of pathogenic strains like *Staphylococcus* aureus underscores the importance of monitoring and addressing AMR in clinically significant pathogens. Additionally, the prevalence of AMR in non-pathogenic strains emphasises the challenge posed by resistance in various staphylococcal species and underscores the need for comprehensive strategies to address antimicrobial resistance.

The examination of antimicrobial agents reveals varying resistance patterns, with some antibiotics demonstrating high rates of resistance. Cloxacillin, for example, showed the highest rates of both AMR and MDR, indicating prevalent resistance within the studied population. Conversely, Ceftazidime exhibited lower rates of resistance, suggesting it may remain effective against certain bacterial isolates.

Finally, the analysis of MDR predictors highlights specimen source and seasonal as important factors to be considered by clinicians when deciding on treatment options.

Overall, findings of the present study, underscore the complex nature of antimicrobial resistance and the need for multifaceted approaches to mitigate its impact on public health. Continued surveillance, prudent antibiotic use, and targeted interventions are essential to combatting antimicrobial resistance effectively.

## Recommendations

Based on the results of the present study on antimicrobial resistance (AMR) in *Staphylococcus* isolates in South Africa, the following recommendations can be made:

- Given the observed higher number of AMR isolates among males compared to females, further research should be conducted to understand the sex-based differences in the risk factors associated with acquiring and transmitting resistant bacterial strains. This could involve exploring potential biological, social, and behavioural factors contributing to these disparities.
- While findings aligned with previous investigations in similar regions, disparities observed in other studies highlight the importance of considering regional differences in AMR prevalence and distribution patterns. Future research should focus on elucidating these regional variations to tailor interventions effectively.
- Given the varying resistance patterns observed among different antimicrobial agents, the prudent use of antibiotics to minimise the development and spread of resistance should be promoted.
- Clinicians should consider local resistance patterns when prescribing antibiotics and prioritize the use of agents with lower rates of resistance, such as Ceftazidime, where appropriate.
- When assessing resistance patterns, specimen source and seasonal variations should be considered. The strong association between blood specimens and MDR occurrence, which may warrant targeted interventions in bloodstream infections, should be taken into consideration.
- Additionally, the seasonal factors influencing resistance patterns to inform seasonal variations in antimicrobial stewardship strategies should be investigated.

By implementing these recommendations, stakeholders can work towards mitigating the impact of antimicrobial resistance on public health and ensuring the continued efficacy of antimicrobial treatments.

## Author Contributions

Conceptualisation, T.T.S., J.W.O. and D.N.Q.; formal analysis, T.T.S. and J.W.O.; data acquisition, T.S.; data curation, T.S.; writing—original draft preparation, T.S.; writing—review and editing, T.S., J.O. and D.Q.; supervision, J.O. and D.Q. All authors have read and agreed to the published version of the manuscript.

## Institutional Review Board Statement

The researcher obtained de-identified data from the diagnostic laboratories, ensuring confidentiality and anonymity by consistently excluding patient information from publications and reports. Additionally, authorisation to use the data was obtained from NHLS. Ethical approval for the study was granted by the University of South Africa, College of Agriculture and Environmental Sciences’ Health Research and Animal Research Ethics Committees (Reference: 2018/CAES/107). Measures were taken to securely protect the data against unauthorised access, accidental loss, or destruction. The data were stored as encrypted files on computers and flash drives in digital format.

## Informed Consent Statement

This is a retrospective study, and it involved accessing de-identified secondary laboratory data containing information on *Staphylococcus* species isolates. No additional procedures or interventions were performed on patients as part of this study. All information obtained from NHLS was kept confidential and was only accessible to authorised members of the research team, and no personal identifying information was included in the analysis or dissemination of results.

## Data Availability Statement

The data that support the findings of this study are available upon reasonable request and under specific conditions. For inquiries regarding access to the data, including requests for collaboration or data sharing agreements, please contact Thomas Papo, Data analyst, at. Requests are considered on a case-by-case basis, taking into consideration the nature of the request, compliance with relevant ethics, and any associated agreements or protocols.

## Acknowledgments

The authors would like to thank the NHLS for giving permission to conduct the study and for providing access to the records used in this study. We also acknowledge the NHLS CDW team, particularly the data analyst, Thomas Papo, for extracting and sharing the *Staphylococcus* data for the 2012-2017 study period.

